# Blood-based epigenome-wide analyses of cognitive abilities

**DOI:** 10.1101/2021.05.24.21257698

**Authors:** Daniel L. McCartney, Robert F. Hillary, Eleanor L. S. Conole, Daniel Trejo Banos, Danni A. Gadd, Rosie M. Walker, Cliff Nangle, Robin Flaig, Archie Campbell, Alison D. D. Murray, Susana Muñoz Maniega, María del. C Valdés-Hernández, Mathew A. Harris, Mark E. Bastin, Joanna M. Wardlaw, Sarah E. Harris, David J. Porteous, Elliot M. Tucker-Drob, Andrew M. McIntosh, Kathryn L. Evans, Ian J. Deary, Simon R. Cox, Matthew R. Robinson, Riccardo E. Marioni

## Abstract

Using blood-based epigenome-wide analyses of general cognitive function (*g*; n=9,162) we show that individual differences in DNA methylation (DNAm) explain 35.0% of the variance in *g*. A DNAm predictor explains ∼4% of the variance in *g*, independently of a polygenic score, in two external cohorts. It also associates with circulating levels of neurology- and inflammation-related proteins, global brain imaging metrics, and regional cortical volumes. As sample sizes increase, our ability to assess cognitive function from DNAm data may be informative in settings where cognitive testing is unreliable or unavailable.

## Manuscript

Blood-based markers of cognitive functioning might provide an accessible way to track neurodegeneration years prior to clinical manifestation of cognitive impairment and dementia. They might also form an easy, objective, and less stressful way to assess neurodegeneration compared to pen-and-paper cognitive tests or in circumstances where biosamples alone are available. Furthermore, they could help to inform our understanding of the biological basis of brain health differences. Blood-based DNA methylation can be used to generate predictors of lifestyle factors, such as smoking, alcohol consumption, and obesity [1] – factors that are linked with poorer cognitive function and an increased risk of dementia [2]. However, blood-based DNA methylation predictors of cognitive function itself, rather than its known correlates, may index a wider range of risk factors for neurodegeneration. Despite being peripheral to the Central Nervous System, blood is an easily accessible tissue, and its DNA methylation patterns may enable early diagnosis and provide mechanistic insights of early phases of disease progression. Although DNA methylation in brain tissue may provide more direct insights into the biology of neurodegeneration, [3, 4] acquiring invivo brain tissue samples is not feasible outside of extraordinary circumstances. Recent methodological advances [5, 6] have enabled the estimation of variance that DNA methylation can account for in complex traits. Therefore, we can now quantify how well blood-based DNA methylation predicts cognitive test outcomes.

There is modest evidence for associations between individual blood-based methylation sites and cognitive functioning; six CpG probes were identified as genome-wide significant in meta-analysis epigenome-wide association studies (EWASs) of seven cognitive traits [7]. That study was limited by heterogeneous cognitive outcomes across cohorts, which also varied in age and ethnicity (meta-analysis n ranging from 2,557 to 6,809). Large-scale single cohort studies with consistent cognitive phenotyping and DNA methylation typing and quality control are lacking. Here, we overcome these limitations by utilising phenotypic cognitive data and blood-based methylation data from a single large cohort of European ancestries.

Blood-based DNA methylation was assessed in 9,162 adult participants from the Generation Scotland cohort [8, 9]. General cognitive ability (*g*) was defined as the first unrotated principal component of tests assessing across four domains: verbal declarative memory (Wechsler Logical Memory test); processing speed (Wechsler Digit Symbol test); executive function (phonemic verbal fluency); and verbal crystallised ability (using Mill Hill Vocabulary synonyms test) [10].

Variance components analyses and epigenome-wide association studies (EWAS) were conducted using BayesR+ software (**Online Methods**). The cognitive phenotypes were corrected for age, sex, BMI and epigenetic smoking score [11], and the DNA methylation data were corrected for batch, age, sex and epigenetic smoking.

A summary of cohort demographics and the phenotypes assessed is presented in **Supplementary Table 1 and Supplementary Figure 1**. The study cohort comprised 59% females and had a mean age of 49.8 years (SD 13.6; range 18 – 93).

Variance components analyses indicated that 41.6% [95% Credible Interval 31.0%, 53.0%] of variance in *g* was explained by all DNA methylation probes (**Supplementary Table 2**). A sensitivity analysis using a linear mixed model approach [5] with genetic and epigenetic relationship matrices yielded near identical results (43.4% (SE 0.03)); a sensitivity analysis using data from an unrelated subset (n=4,261) of the study cohort that was processed in a single methylation batch also showed similar estimates (58.4% (SE 0.07); **Supplementary Table 3**). The majority of the variance was accounted for by CpG sites with small effects; three prior mixture distributions were set with small (0.01%), medium (0.1%), and large (1%) variances for effect sizes (**Supplementary Table 4**).

The SNP-based heritability was 37.9% [18.3%, 25.9%], which is in line with previous GREML estimates from the cohort [12]. The proportion of variance explained when combining the independent effects of genetics and DNAm increased to 63.8% [50.0%, 73.5%] (**Supplementary Table 2**). Notably, the CpG contribution to the variance accounted for was largely independent of the genetic component – absolute attenuation 6.6% (relative attenuation 15.9%) to the epigenetic effect size estimate in the model that included genetics.

The EWAS model identified three unique lead DNAm sites with a posterior inclusion probability (PIP) greater than 0.80 and a group-based PIP>0.95 (**Supplementary Figure 2; Supplementary Table 5;** the entire output is available at https://gitlab.com/danielmccartney/ewas_of_cognitive_function). For the three lead CpG sites we queried the EWAS catalog (accessed 5 April 2021) for associations with other traits (**Supplementary Table 6**) [13]. They have been linked to age, metabolite levels, lipid levels, kidney cancer, and lung function. Of 28 CpGs identified in previous blood-based EWAS analyses of cognitive ability and Alzheimer’s disease [4, 7, 14], 24 were available for lookup in the present dataset; Generation Scotland data was not included in any of these studies. There was no evidence for replication (maximum PIP of 0.03; **Supplementary Table 7**).

We applied the mean posterior effect sizes for all possible CpGs from the EWAS model of *g* to CpGs in two independent studies (The Lothian Birth Cohort 1936 (LBC1936) and The Lothian Birth Cohort 1921 (LBC1921), n=844 and n=427 with DNAm, cognitive scores, and cognitive polygenic score available, respectively – **Online Methods**). The incremental R^2^ upon the addition of the Epigenetic Score (EpiScore) to a linear regression model adjusting for age and sex was 3.4% (P=2.0x10^-8^) in LBC1936 and 4.5% (P=9.9x10^-6^) in LBC1921. The corresponding R^2^ for the polygenic score derived from a GWAS of 168,033 individuals in UK Biobank – a training sample over 18-times greater than the DNAm training sample – was 7.3% (P<2x10^-16^) and 6.9% (P=3.1x10^-8^), respectively. The additive incremental R^2^ from the two omics-based predictors was 10.7% and 10.5% (**Figure 1**).

**Figure 1:**
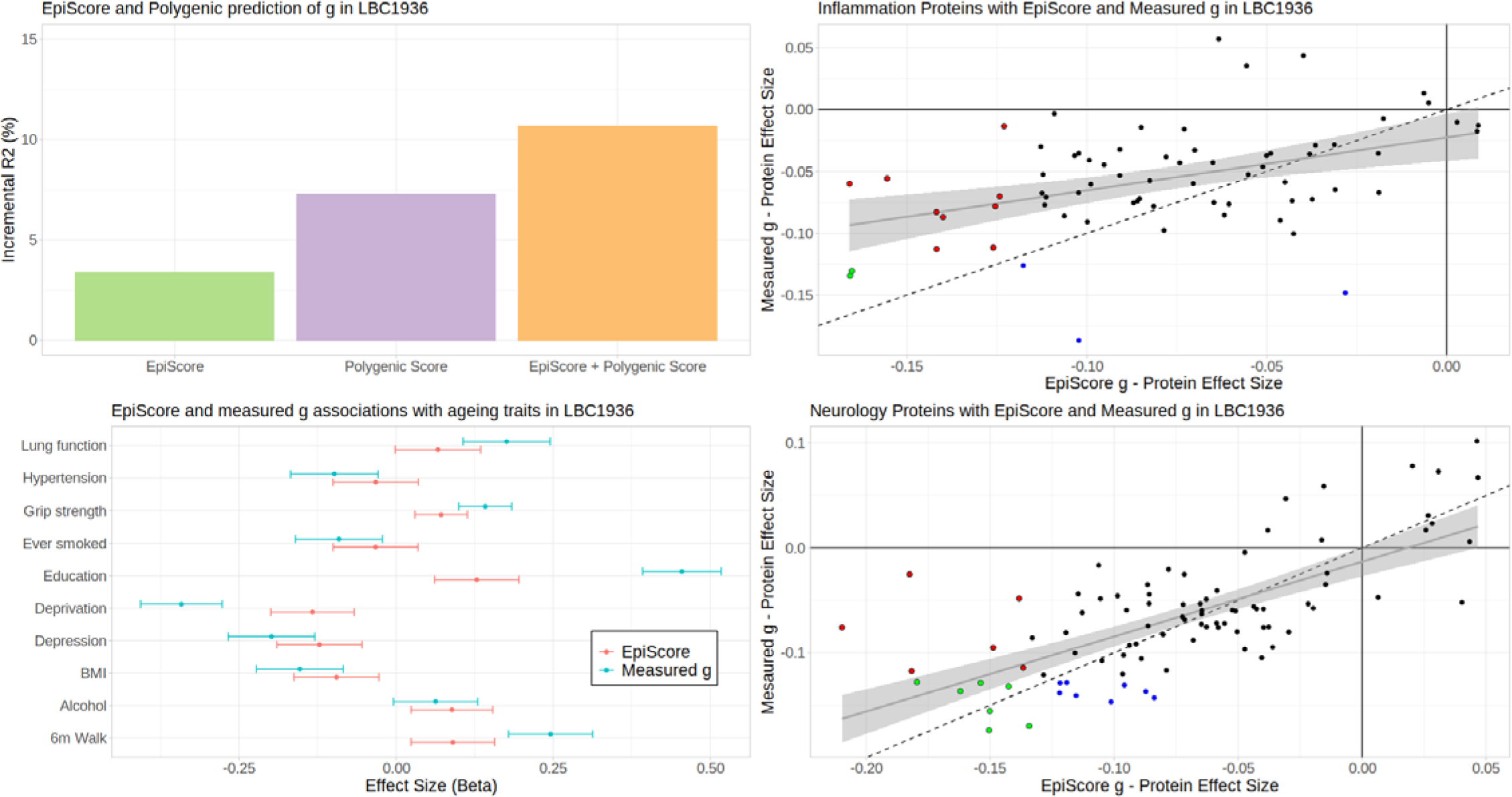
An epigenetic score for cognitive ability associates with measured cognitive ability, health and lifestyle factors, and neuro-inflammatory protein levels. Variance explained for general cognitive ability (*g*) by a cognitive Epigenetic Score (EpiScore; green), polygenic score (purple), and in combination (orange) (A). Age- and sex-adjusted associations between risk factors for cognitive decline and dementia and the EpiScore (red) and measured *g* (turquoise) with 95% confidence intervals – deprivation and 6m walk have been reverse coded such that higher values correspond to less deprivation and faster walking speed (B). Comparison of age- and sex-adjusted associations between the EpiScore and measured g score with 70 inflammation-related (C) and 90 neurology related (D) proteins. Coloured points in C and D are significant after Bonferroni-correction: green – common to both, red – unique to EpiScore, blue – unique to measured g; dashed lines show perfect correlation (y=x) the grey lines show the linear regression slope with 95% confidence interval.

Based on a training sample size of 10,000 and assuming 100,000 CpGs affect the trait, with a DNAm variance components estimate of 41.6%, we would expect a DNAm prediction R^2^ value of about 4.0% (following Formula 1 from [15]). This is very similar to the estimates obtained. If the training sample increased to 20,000 or 100,000 then the expected R^2^ should increase to around 8% and 29%, respectively.

In age- and sex-adjusted linear regression analyses with common risk factors of cognitive decline (smoking, years of education, BMI, lung function, walking speed, grip strength, high blood pressure, alcohol consumption, a depression questionnaire score, and an index of social deprivation), the EpiScore showed directionally consistent but weaker associations than measured *g* across both LBC1921 and LBC1936 (**Figure 1** **and Supplementary Table 8**). The only exception was alcohol consumption, where the EpiScore outperformed measured *g*.

Age- and sex-adjusted linear regression analyses were conducted with 70 Olink inflammatory protein levels in LBC1936. The EpiScore and measured *g* associations with the proteins were moderately concordant (r = 0.43). The EpiScore associated with 11 proteins (P<0.05/70) compared to 5 for measured *g* with two proteins, IL-6 and FGF21, overlapping both sets (**Figure 1** **and Supplementary Table 9**).

DNA methylation, structural brain MRI, and 90 Olink neurology-related proteins were also available at a follow-up wave of LBC1936 when participants were a mean age of 73 years (n=701 with proteins and n=551 with MRI). The EpiScore – protein associations mirrored those previously reported with measured fluid cognitive ability in the same dataset [16] with similar effect size estimates (r = 0.70). Thirteen EpiScore- and 15 measured *g*-protein associations were statistically significant (P<0.05/90) with 7 overlapping (**Figure 1** **and Supplementary Table 9**). There were associations with brain imaging measures of global volume (total brain, grey matter, and normal appearing white matter volumes – **Figure 2** and **Supplementary Table 10**). Furthermore, there were widespread associations between the EpiScore and regional brain cortical volume and thickness (**Figure 2** and **Supplementary Figures 4-5**), with significant overlap in cortical loci for both measured g and EpiScore. Overall, the EpiScore findings largely mirrored the associations between measured g and neuroimaging outcomes, albeit they were slightly smaller in magnitude.

**Figure 2.**
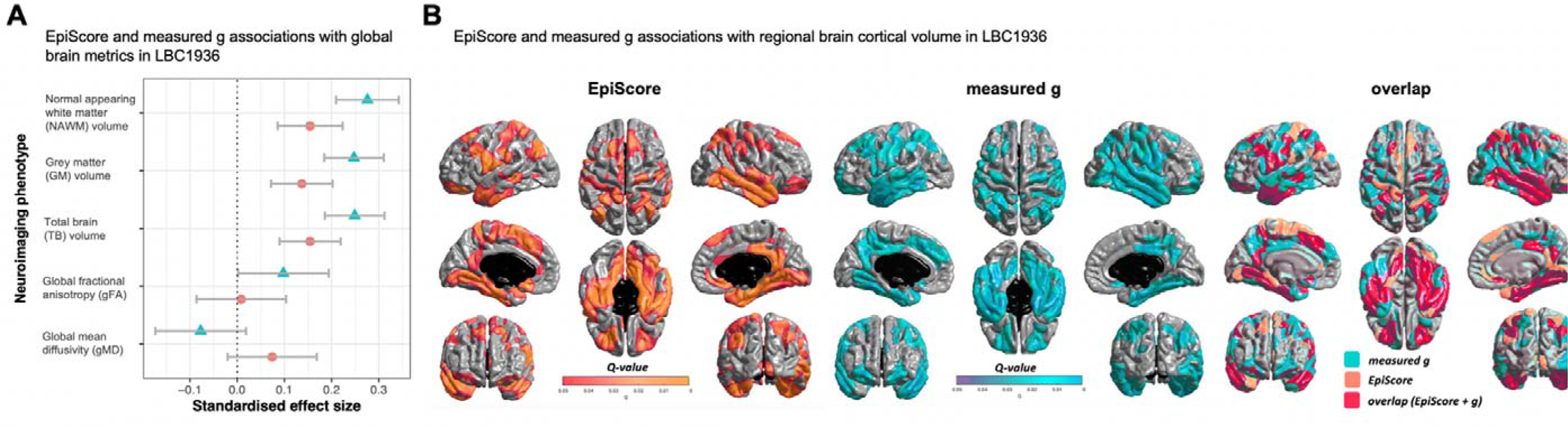
Measured and epigenetic cognitive ability associate with brain structure and show regional overlap with cortical loci. (A) Cognitive ability measures with global brain imaging associations in LBC1936 with 95% confidence intervals; measured *g* (turquoise), Epigenetic Score (EpiScore; orange). (B) Results of cortical volume at age 73 years regressed against cognitive *g* EpiScore (orange), measured *g* (turquoise) and the spatial extent of overlap (pink) in cortical loci. Colours, representing q values, are superimposed on an average surface template. A false discovery rate threshold of 0.05 is used to control for multiple comparisons; results are corrected for sex, age in days at brain scanning and intracranial volume (n=551)

This is the first variance components analysis of DNAm and cognitive function. We show substantial contributions to several of the tests in addition to the development of a novel epigenetic score with application to two independent test cohorts where it associated with cognitive ability. We also show associations between the EpiScore and lifestyle factors and risk factors for dementia, circulating levels of neuroinflammatory proteins, and brain MRI measures.

We used a large homogeneous discovery cohort with consistent cognitive testing and DNA collection across all participants. This will minimise the biases that are an inherent problem in heterogeneous and small EWAS meta-analyses. The estimation of posterior effect sizes using BayesR+ has been shown to be robust to various potential sources of heterogeneity, including family structure [17] and population structure based on genetic or epigenetic principal components [6]. Nonetheless, replication of our findings in cohorts of different ages and backgrounds will help to refine and generalise the estimates presented here.

Methylation-based predictors of cognitive function may improve longitudinal disease prediction and risk profiling of neurodegenerative health outcomes such as dementia. Here, the epigenetic variance components and prediction score were independent of genetic contributions; the EpiScore also reflected measured cognitive ability in its associations with a variety of biological and health-based traits. Unlike DNA differences which are largely fixed throughout life, DNAm differences may reflect environmental effects and phenotypic causation, directly through cognitive-related pathways or indirectly via related lifestyle and health outcomes. Given the overwhelming contribution of CpGs with small effects to our estimates, increasing EWAS sample sizes and employing powerful statistical methods like BayesR+ will clearly lead to both locus discovery and more accurate DNAm-based predictors of cognitive function. Whereas the associations of the current EpiScore with lifestyle and MRI variables are more modest than those observed for measured *g*, this gap is likely to narrow as the sample size of the training set increases. This has potential implications for studies of cognitive function across the lifespan where pen-and-paper testing is not possible or unreliable, such as during neurodevelopment or neurodegeneration.

## Declarations

### Ethics approval and consent to participate

All components of GS received ethical approval from the NHS Tayside Committee on Medical Research Ethics (REC Reference Number: 05/S1401/89). GS has also been granted Research Tissue Bank status by the East of Scotland Research Ethics Service (REC Reference Number: 20-ES-0021), providing generic ethical approval for a wide range of uses within medical research.

Ethical approval for the LBC1921 and LBC1936 studies was obtained from the Multi-Centre Research Ethics Committee for Scotland (MREC/01/0/56) and the Lothian Research Ethics committee (LREC/1998/4/183; LREC/2003/2/29). In both studies, all participants provided written informed consent. These studies were performed in accordance with the Helsinki declaration.

### Availability of Data and Material

According to the terms of consent for GS participants, access to data must be reviewed by the GS Access Committee. Applications should be made to.

Lothian Birth Cohort data are available on request from the Lothian Birth Cohort Study, University of Edinburgh. Lothian Birth Cohort data are not publicly available due to them containing information that could compromise participant consent and confidentiality.

All code is available with open access at the following Gitlab repository: https://gitlab.com/danielmccartney/ewas_of_cognitive_function

## Data Availability

According to the terms of consent for GS participants, access to data must be reviewed by the GS Access Committee. Applications should be made to.
All code is available with open access at the following Gitlab repository: https://gitlab.com/danielmccartney/ewas_of_cognitive_function

## Acknowledgements

GS received core support from the Chief Scientist Office of the Scottish Government Health Directorates (CZD/16/6) and the Scottish Funding Council (HR03006). Genotyping and DNA methylation profiling of the GS samples was carried out by the Genetics Core Laboratory at the Edinburgh Clinical Research Facility, Edinburgh, Scotland and was funded by the Medical Research Council UK and the Wellcome Trust (Wellcome Trust Strategic Award STratifying Resilience and Depression Longitudinally (STRADL; Reference 104036/Z/14/Z). The DNA methylation data assayed for Generation Scotland was partially funded by a 2018 NARSAD Young Investigator Grant from the Brain & Behavior Research Foundation (Ref: 27404; awardee: Dr David M Howard) and by a JMAS SIM fellowship from the Royal College of Physicians of Edinburgh (Awardee: Dr Heather C Whalley). LBC1936 MRI brain imaging was supported by Medical Research Council (MRC) grants [G0701120], [G1001245], [MR/M013111/1] and [MR/R024065/1]. Magnetic Resonance Image acquisition and analyses were conducted at the Brain Research Imaging Centre, Neuroimaging Sciences, University of Edinburgh (www.bric.ed.ac.uk) which is part of SINAPSE (Scottish Imaging Network: A Platform for Scientific Excellence) collaboration (www.sinapse.ac.uk) funded by the Scottish Funding Council and the Chief Scientist Office. This work was supported by the Centre for Cognitive Ageing and Cognitive Epidemiology, funded by the Medical Research Council and the Biotechnology and Biological Sciences Research Council (MR/K026992/1), the Row Fogo Charitable Trust (BRO.D.FID3668413), the European Union Horizon 2020, (PHC.03.15, project No 666881), SVDs@Target, the Fondation Leducq Transatlantic Network of Excellence for the Study of Perivascular Spaces in Small Vessel Disease [ref no. 16 CVD 05], and the Medical Research Council UK Dementia Research Institute at the University of Edinburgh. We thank the LBC1936 participants and team members who contributed to these studies. The LBC1936 is supported by Age UK (Disconnected Mind project, which supports S.E.H.), the Medical Research Council (G0701120, G1001245, MR/M013111/1, MR/R024065/1), and the University of Edinburgh. Methylation typing of LBC1936 was supported by the Centre for Cognitive Ageing and Cognitive Epidemiology (Pilot Fund award), Age UK, The Wellcome Trust Institutional Strategic Support Fund, The University of Edinburgh, and The University of Queensland. Genotyping was funded by the Biotechnology and Biological Sciences Research Council (BB/F019394/1). Proteomic analyses in LBC1936 were supported by the Age UK grant and NIH Grants R01AG054628 and R01AG05462802S1. M.V.H. is funded by the Row Fogo Charitable Trust (Grant no. BROD.FID3668413). J.M.W is supported by the UK Dementia Research Institute which receives its funding from DRI Ltd, funded by the UK Medical Research Council, Alzheimers Society and Alzheimers Research UK. R.F.H., E.L.S.C and D.A.G. are supported by funding from the Wellcome Trust 4 year PhD in Translational Neuroscience: training the next generation of basic neuroscientists to embrace clinical research [108890/Z/15/Z]. E.M.T.D. was supported by National Institutes of Health (NIH) grants R01AG054628, R01MH120219, R01HD083613, P2CHD042849 and P30AG066614. S.R.C. was also supported by a National Institutes of Health (NIH) research grant R01AG054628, and is supported by a Sir Henry Dale Fellowship jointly funded by the Wellcome Trust and the Royal Society (Grant Number 221890/Z/20/Z). D.L.Mc.C. and R.E.M. are supported by Alzheimers Research UK major project grant ARUK/PG2017B/10. This research was funded in whole, or in part, by Wellcome [104036/Z/14/Z and 108890/Z/15/Z]. For the purpose of open access, the author has applied a CC BY public copyright licence to any Author Accepted Manuscript version arising from this submission.

## Online Methods

### The Generation Scotland Cohort

Details of the Generation Scotland: Scottish Family Health Study (GS) have been described in detail elsewhere [9, 10]. Briefly, GS comprises over 20,000 individuals comprehensively profiled for genetic, clinical, lifestyle, and sociodemographic data. A subset of 9,162 individuals from GS (aged 18 to 93 years, mean=49.7, SD=13.6) had genome-wide DNA methylation measured [18]. This subset was processed in two batches, hereafter referred to as “Set 1” and “Set 2”.

### Methylation preparation in Generation Scotland

Quality control was performed on Illumina HumanMethylationEPIC BeadChip DNA methylation data from blood samples of 5,200 related individuals from Set 1, and 4,588 genetically unrelated individuals from Set 2, also genetically unrelated to those in Set 1. Three Set 1 individuals who had answered “yes” to presence of all of 16 self-reported disease conditions in the study’s health questionnaire were excluded from the analysis. Filtering for outliers, sex mismatches, non-blood samples, and poorly detected probes and samples was performed [18]. Further filtering was then carried out to remove CpGs with missing values, non-autosomal and non-CpG sites. Five individuals with a self-reported diagnosis of Alzheimer’s disease were removed, along with samples with missing covariate information or cognitive variables. Following sample filtering, 9,162 complete cases remained comprising 4,901 Set 1 individuals and 4,261 Set 2 individuals.

### Cognitive Phenotypes in Generation Scotland

Six cognitive phenotypes were assessed in this study: logical memory, digit symbol test score, verbal fluency, vocabulary, general cognitive ability, and general fluid cognitive ability. The logical memory phenotype (verbal declarative memory) was calculated from the Wechsler Logical Memory test, taking the sum of immediate and delayed recall of one oral story [19]. The digit symbol phenotype is often used as a measure of processing speed; calculated from the Wechsler Digit Symbol Substitution test in which participants must recode digits to symbols over a 120 second period [20]. The verbal fluency phenotype is often used as a measure of executive functioning and was derived from the phonemic verbal fluency test, using the letters C, F, and L, each for one minute [21]. Vocabulary was measured using the Mill Hill Vocabulary Scale, junior and senior synonyms combined [22]. General fluid cognitive ability (*g_f_*) was calculated from the first unrotated principal component of logical memory, verbal fluency, and digit symbol tests. General cognitive ability (*g*) was derived from the first unrotated principal component from the same variables plus vocabulary.

### Statistical Analysis

BayesR+ was also used for the EWAS and to estimate the genetic and DNA methylation-based heritability of the cognitive traits. BayesR+ is a software implemented in C++ for performing Bayesian penalised regression and Gaussian mixture-based variance partitioning on complex traits [6]. The joint and conditional effects of methylation sites (n=764,525) on cognitive traits were examined. Phenotypic and methylation data were scaled to mean zero and unit variance. The prior distribution comprised a series of Gaussian distributions which corresponded to effect sizes of different magnitudes (i.e. methylation sites with small, medium and large effect sizes), as well as a discrete spike at zero which allows for the omission of probes with non-identifiable effects. The prior mixture variances were set to 0.0001, 0.001 and 0.01. Phenotypes were corrected for age, sex, BMI and epigenetic smoking score [11], and the DNA methylation data were corrected for batch, age, sex and epigenetic smoking. Adding a quadratic term for age in the correction of cognitive phenotypes made negligible differences to the trait residuals (r>0.98 between linear model residuals for each trait). To obtain estimates of variance accounted for in cognitive traits by methylation data and individual CpG associations with the cognitive test scores, Gibbs sampling was performed to sample over the posterior distribution conditional on the input data. The Gibbs algorithm consisted of 10,000 samples and 5,000 samples of burn-in after which a thinning of 5 samples was applied to reduce autocorrelation. The process was repeated over four chains, initializing a different random number seed for each chain. The last 250 iterations from each chain were combined for downstream analyses. For the EWAS, CpGs within 2.5kb and highly correlated (absolute Pearson correlation >0.5) with a lead CpG with posterior inclusion probability greater than 0.2 were grouped together. For each probe group, we calculated the proportion of iterations for which at least one probe was included in the model, yielding the group posterior inclusion probability. We then calculated the average (across the 1,000 iterations) sum of the squared regression coefficients for the probe group to give the contribution of the group to the total variance. Finally, we highlighted the lead CpG for the groups where the combined posterior inclusion probability was >0.80. Genetic effects at 560,797 SNPs (minor allele frequency > 1%; scaled to mean 0, variance 1) from the Illumina HumanOmniExpressExome-8 v1.0 Bead Chip or Illumina HumanOmniExpressExome-8 v1.2 Bead Chip were examined [23], setting prior mixture variances to 0.00001, 0.0001 and 0.001. For the genetic analysis, phenotypes were pre-corrected for age, sex and 20 genetic PCs. To estimate the additive and independent effects of DNAm and genetic data on complex traits, a combined analysis was run – using the phenotype corrections specified for the EWAS model setting prior mixture variances as above.

The mean posterior effect sizes for the EWAS model of general cognitive function, g, were used to generate an epigenetic predictor in two independent datasets, the Lothian Birth Cohort 1936 (LBC1936) and the Lothian Birth Cohort 1921 (LBC1921). The LBC are studies of cognitive ageing in older adults from the area around Edinburgh, Scotland [24, 25]. Briefly, participants were born in either 1921 or 1936 and completed the Scottish Mental Survey of 1932 or1947 at age 11. From age 70 (LBC1936) and age 79 (LBC1921), they were assessed triennially for a variety of health and lifestyle outcomes, with DNA collected at each visit. Here, we considered blood-based DNA methylation data from the age 70 (LBC1936) and age 79 (LBC1921) samples. Methylation was assessed on the Illumina 450k array – the predecessor of the EPIC array. Processing and quality control have been described previously [1, 26, 27]. This included steps to remove methylation samples and individuals with poor quality control measures, along with individuals who had mismatching genotypes or predicted sex information. This left a dataset of 381,846 CpGs (overlapping with those included in the Generation Scotland analyses) for 861 LBC1936 (436 LBC1921) individuals 34 LBC1936 individuals were excluded due to DNAm being assessed as part of a separate analysis batch. Within each cohort, each CpG was scaled to mean 0, variance 1 with missing values mean imputed (i.e., set to 0) prior to multiplication by the mean CpG weights (for all available CpGs) and summation to give the epigenetic score. A polygenic score for cognitive ability was derived from Z-scores from all possible SNPs (GWAS P<1) in a UK Biobank GWAS of verbal numerical reasoning (n=168,033) [30] and applied to LBC1936 and LBC1921 genotype data using default settings in the PRSice software [31, 32].In LBC1936, general cognitive ability was defined as the first unrotated principal component (that accounted for 52% of the variance) from a PCA of six cognitive tests from the Wechsler Adult Intelligence Scale-III UK (matrix reasoning, letter number sequencing, block design, symbol search, digit symbol, and digit span backward) [20] plus a test of vocabulary (National Adult Reading Test) [28]. A similar approach was taken in LBC1921, where we considered the first unrotated principal component (that also accounted for 51% of the variance) from a PCA of four cognitive tests: Raven’s Standard Progressive Matrices [29], letter-number sequencing [20], digit symbol coding [20], and the National Adult Reading Test) [28]. The cognitive tests were completed at the same visit that blood was drawn for DNA profiling in both LBC studies. There were 844 LBC1936 (427 LBC1921) individuals with cognitive, epigenetic, and polygenic score data. Linear regression was used to test for an association between the predicted epigenetic score (predictor) and measured general cognitive ability (outcome), in models adjusting for age, sex, and the polygenic score. Age- and sex-adjusted linear regression models were used to relate the Epigenetic Score (EpiScore) for *g* and the measured *g* score (predictors) with common risk factors (outcomes) for cognitive decline, frailty, and dementia: body mass index (BMI in kg/m^2^); year of education; self-reported smoking (ever versus never); self-reported weekly units of alcohol; self-reported high blood pressure (yes/no); lung function (forced expiratory volume in one second) adjusted for age, sex, and height; time taken to walk six metres (seconds); socioeconomic deprivation (Scottish Index of Multiple Deprivation in LBC1936 and social grades based on highest reached occupation [33] in LBC1921); and depression (HADS-D total from the Hospital Anxiety and Depression questionnaire) [34, 35, 36]. BMI, six metre walk time, and units of alcohol were log transformed to reduce skew – a constant of one was added to the alcohol units prior to transforming. An Olink panel of inflammation-related proteins [37], measured on blood samples at age 70 years in LBC1936, were related to both EpiScore *g* and measured *g* in age- and sex-adjusted linear regression models. An additional panel of Olink neurology-related proteins [16, 37], measured on blood samples at age 73 years in LBC1936, were related to EpiScore *g*, which was derived from DNAm assessed from the same sample (analysis n=701). Quality control of the DNAm was identical to the age 70 samples. Processing occurred in two sets (N_Set1_=256, N_Set2_=445) where CpG sites were independently scaled to mean zero and variance one, prior to combining into a single variable. Each of the 70 inflammatory and 90 neurology proteins were adjusted via rank-based inverse normal transformations and regressed on age, sex, and four genetic ancestry components as previously described [38]. Linear regression model assumptions were visually inspected via regression diagnostic plots. Structural and diffusion tensor (DTI) MRI acquisition and processing in LBC1936 were performed at Wave 2 (age 73 years) according to an open-access protocol [39]. A 1.5□T GE Signa HDx clinical scanner (General Electric, Milwaukee, WI, USA) was used to collect structural T1- (voxel size = 1 × 1 × 1.3 mm), T2- (voxel size = 1 × 1 × 2 mm, T2*- (voxel size = 1 × 1 × 2 mm), and FLAIR-weighted images (voxel size = 1 × 1 × 4 mm). Diffusion MRI protocol consisted of a single-shot spin-echo echo-planar diffusion-weighted sequence. Diffusion-weighted volumes (b = 1000 s mm−2) were acquired in 64 non-collinear directions, with seven T2-weighted volumes (b□=□0 s mm−2), with 72 contiguous axial slices, and an acquisition matrix of 128□×□128 and 2-mm isotropic voxels. Total brain, grey matter, and normal-appearing white matter (NAWM) volumes were calculated using a semi-automated multi-spectral fusion method [40]. Intracranial volume was determined semi-automatically using Analyze 11.0^TM^. White matter microstructural parameters fractional anisotropy (FA) and mean diffusivity (MD) were derived for 12 major tracts of interest: corpus callosum genu and splenium, bilateral frontal cingulum, arcuate, uncinate and superior longitudinal fasciculi and bilateral anterior thalamic radiation. These were obtained using probabilistic neighbourhood tractography in TractoR (https://www.tractor-mri.org.uk) [41, 42] as applied to BEDPOSTX/ProbTrackX in FSL (https://fsl.fmrib.ox.ac.uk) [43]. The FA and MD values from each of the 12 tracts were the weighted average of the diffusion values contained within the resultant tract map. General measures of FA and MD (gFA and gMD [44]) were the extracted scores from the first unrotated principal component, accounting for 37% and 41% of the variance in tract FA and MD, respectively. Cortical reconstruction and segmentation was performed using FreeSurfer v5.1 on T1-weighted volumes. QC involved visually assessing each image output for segmentation and parcellation errors, which were then corrected manually; segmentations with errors that could not be corrected were excluded. Participants were then excluded if they had self-reported history of dementia or signs of cognitive impairment (Mini mental state examination score < 24/30); after exclusions, a total of 590 participants had complete cognitive, epigenetic and global neuroimaging data, and of these, 551 participants had complete and vertex-wise neuroimaging data. Localized associations between cognitive measures and vertex-wise cortical volume and thickness were performed using linear regression, controlling for age, sex, and Intracranial volume (ICV). The SurfStat MATLAB toolbox (http://www.math.mcgill.ca/keith/surf stat) for Matrix Laboratory R2012a (The MathWorks, Inc., Natick, MA, USA) was used to carry out analyses. Statistical maps of association magnitude and valence (*t*-maps) and significance (q-maps; p-values corrected for multiple comparisons using a false discovery rate (FDR) with a q-value of 0.05 across all 327,684 vertices on the cortical surface) were presented.

---

**Supplementary Table 1:** Generation Scotland cohort summary

|  | N | Mean | SD |
| --- | --- | --- | --- |
| Age (years) | 9162 | 49.8 | 13.6 |
| Digit Symbol | 9162 | 71.2 | 16.5 |
| Verbal Fluency | 9162 | 40.0 | 11.5 |
| Vocabulary | 9162 | 30.5 | 4.5 |
| Logical Memory | 9162 | 31.1 | 7.9 |
| Body Mass Index (kg/m<sup>2</sup>) | 9162 | 26.9 | 5.2 |
| EpiSmoker* | 9162 | 2.09 | 4.5 |
|  | N | % |  |
| Sex |  |  |  |
| Male | 3752 | 41.0 |  |
| Female | 5410 | 59.0 |  |

\* A plot of the epigenetic smoker (EpiSmoker) variable against self-reported smoking (current, former, never) is provided in **Supplementary Figure 1**.

**Supplementary Table 2:**
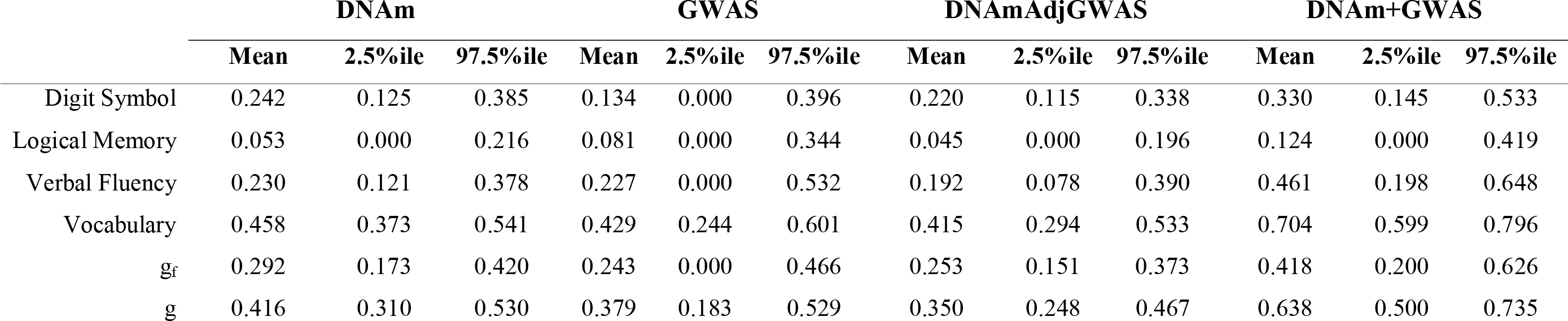
Mean variance accounted for by the effects of DNA methylation (DNAm) and genome-wide DNA single nucleotide polymorphisms (GWAS) alone, DNAm data conditioned on GWAS data (DNAmAdjGWAS), and the additive effects of DNAm and GWAS data for six cognitive traits.

**Supplementary Table 3:** Comparison of epigenetic variance components estimates between BayesR+ and a linear mixed model approach, OSCA. OSCA^1^ analyses included genetic and epigenetics relationship matrices on all 9,162 participants; OSCA^2^ analyses included an epigenetics relationship matrix on a genetically unrelated subset of 4,261 participants who were processed in a single methylation batch (“Set 2” – **Online Methods**).

| DNAm variance component estimate |  |  |  |
| --- | --- | --- | --- |
|  | BayesR+ | OSCA<sup>1</sup> | OSCA<sup>2</sup> |
|  | (mean) | (mean, SE) | (mean, SE) |
| Logical Memory | 0.053 | 0.062 (0.03) | 0.142 (0.08) |
| Digit Symbol | 0.242 | 0.257 (0.04) | 0.231 (0.07) |
| Verbal Fluency | 0.230 | 0.254 (0.04) | 0.168 (0.07) |
| Vocabulary | 0.458 | 0.484 (0.03) | 0.604 (0.06) |
| g<sub>f</sub> | 0.292 | 0.313 (0.04) | 0.417 (0.08) |
| g | 0.416 | 0.434 (0.03) | 0.584 (0.07) |

**Supplementary Table 4:**
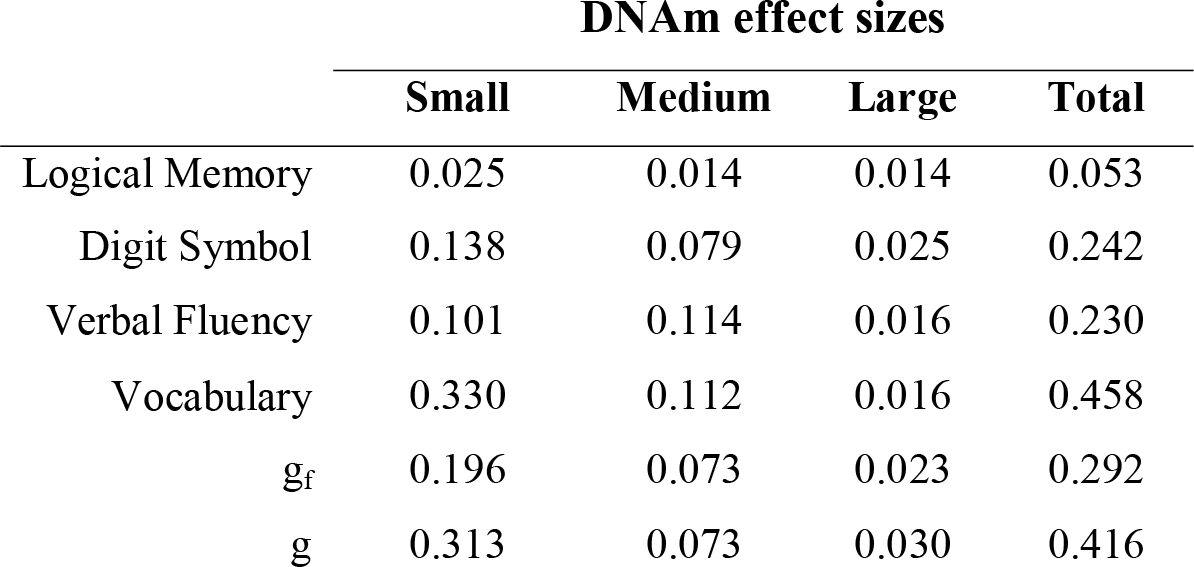
Contribution of mixtures with small, medium, and large effect sizes (variances of 0.01%, 0.1%, and 1%, respectively) to the mean variance accounted for by the effects of DNA methylation (DNAm) for the cognitive traits.

|  | **DNAm effect sizes** |  |  |  |
| --- | --- | --- | --- | --- |
|  | **Small** | **Medium** | **Large** | **Total** |
| Logical Memory | 0.025 | 0.014 | 0.014 | 0.053 |
| Digit Symbol | 0.138 | 0.079 | 0.025 | 0.242 |
| Verbal Fluency | 0.101 | 0.114 | 0.016 | 0.230 |
| Vocabulary | 0.330 | 0.112 | 0.016 | 0.458 |
| g<sub>f</sub> | 0.196 | 0.073 | 0.023 | 0.292 |
| g | 0.313 | 0.073 | 0.030 | 0.416 |

**Supplementary Table 5:** List of blood-based CpG sites that are with strong evidence (Group PIP>0.95) for association with general cognitive function in Generation Scotland. PIP: Posterior Inclusion Probability. For the EWAS, CpGs within 2.5kb and highly correlated (absolute Pearson correlation >0.5) with a lead CpG with posterior inclusion probability greater than 0.2 were grouped together. For each probe group (with number of CpGs – **nCpG**), we calculated the proportion of iterations for which at least one probe was included in the model, yielding the group posterior inclusion probability (**Group PIP**). We then calculated the sum of the squared regression coefficients at each iteration for the probe group to give the contribution of the group to the total variance (**Group R^2^**). Finally, we highlighted the lead CpG for the groups.

| Lead CpG | Group PIP | Group R<sup>2</sup> | Nearest Gene | nCpG | Lead PIP | Lead Median Beta | Lead Beta2.5 | Lead Beta97.5 |
| --- | --- | --- | --- | --- | --- | --- | --- | --- |
| cg09984392 | 1.013 | 0.005 | *SQLE* | 4 | 0.965 | 0.072 | 0.000 | 0.104 |
| cg13108341 | 1.000 | 0.009 | *DNAH9* | 1 | 1.000 | -0.092 | -0.124 | -0.054 |
| cg05325763 | 0.952 | 0.005 | *CPT1A* | 5 | 0.870 | -0.070 | -0.108 | 0.000 |

**Supplementary Table 6:**
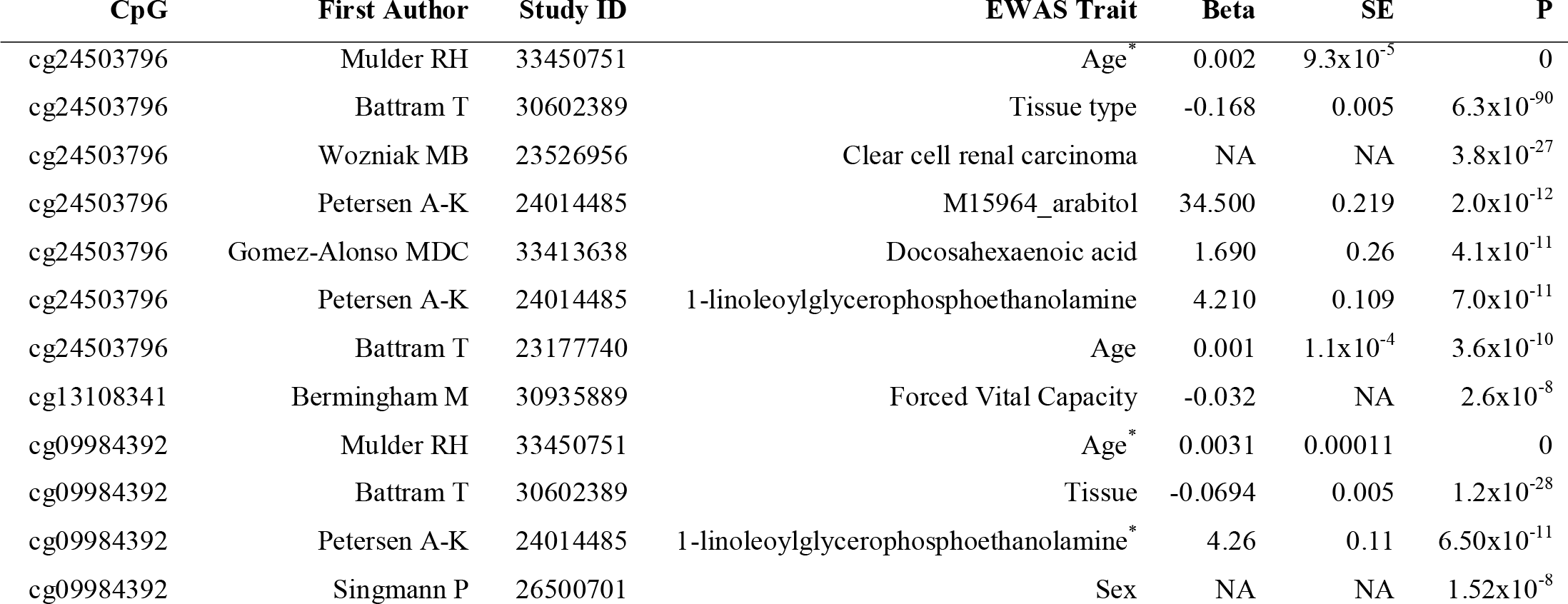
Epigenome-wide association study (EWAS) catalog lookup of three CpGs with strong (Group PIP>0.95) associations with general cognitive ability. Lookup is restricted to associations at P<3.6x10^-8^. * denotes multiple models in study, where summary statistics are presented for the model with the most significant association.

**Supplementary Table 7:**
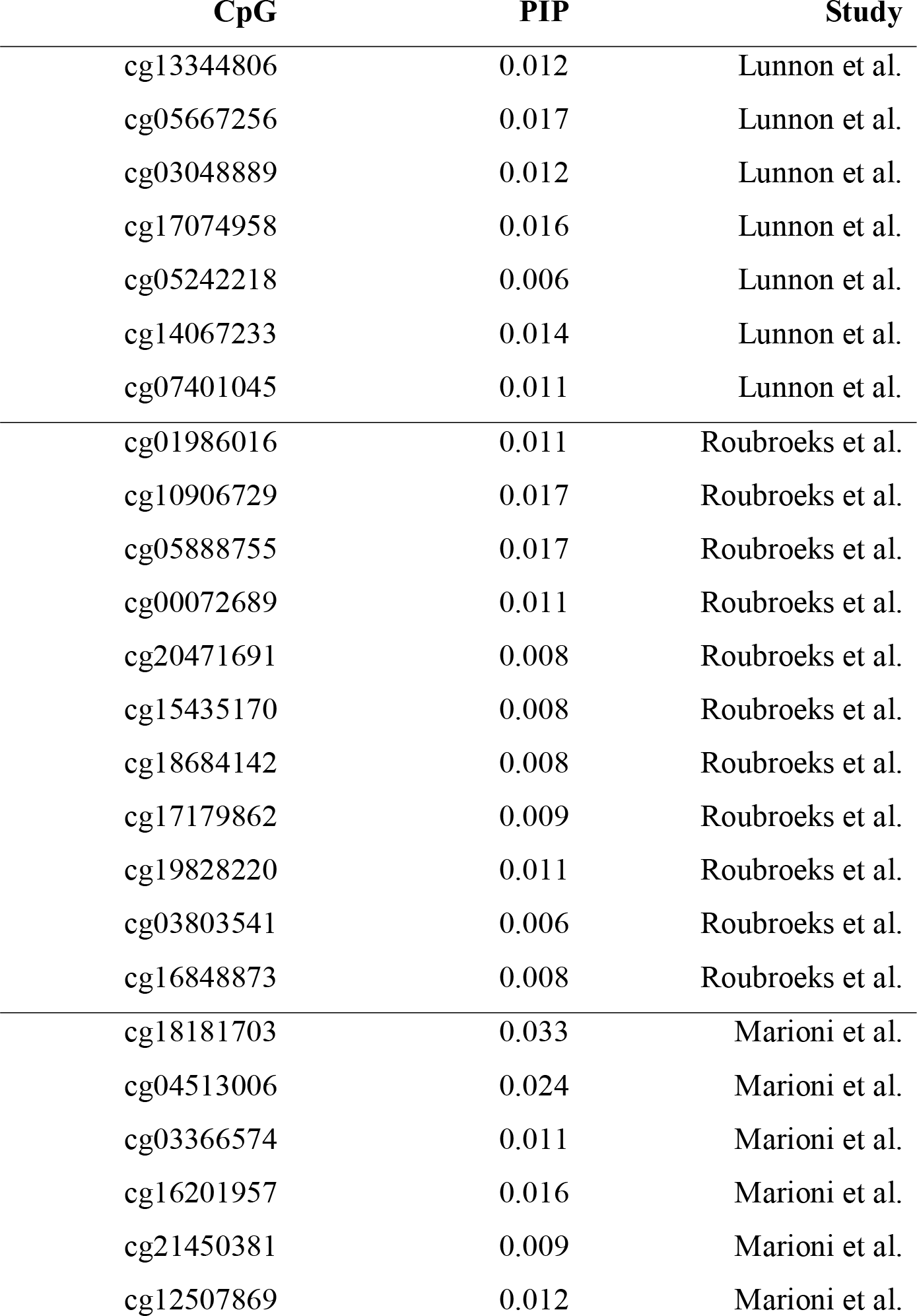
Lookup of CpGs associated with cognitive abilities and Alzheimer’s disease. PIP (Posterior Inclusion Probability) is presented for *g*, as reported in the current study.

**Supplementary Table 8.**
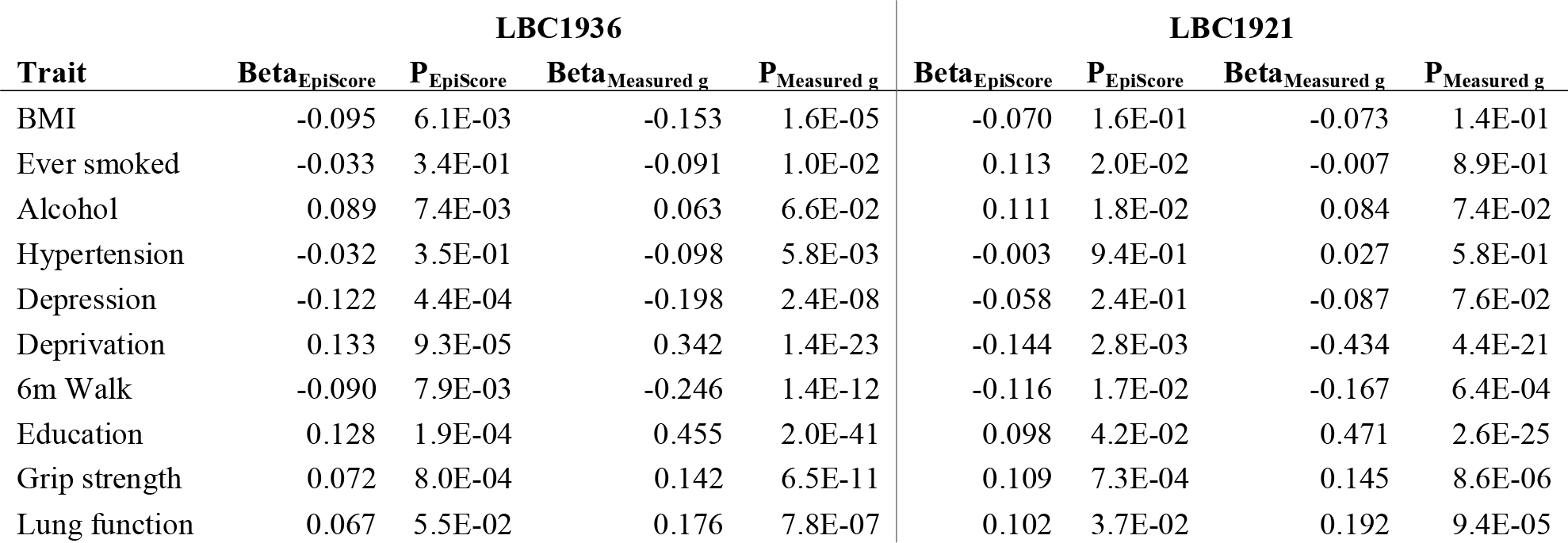
Age- and sex-adjusted linear regression associations between the cognitive *g* Epigenetic Score (EpiScore) and measured *g* score and traits associated with cognitive ageing and dementia (outcomes) in the Lothian Birth Cohorts 1936 and 1921. All continuous variables are standardised to mean 0 and variance 1.

**Supplementary Table 9.**
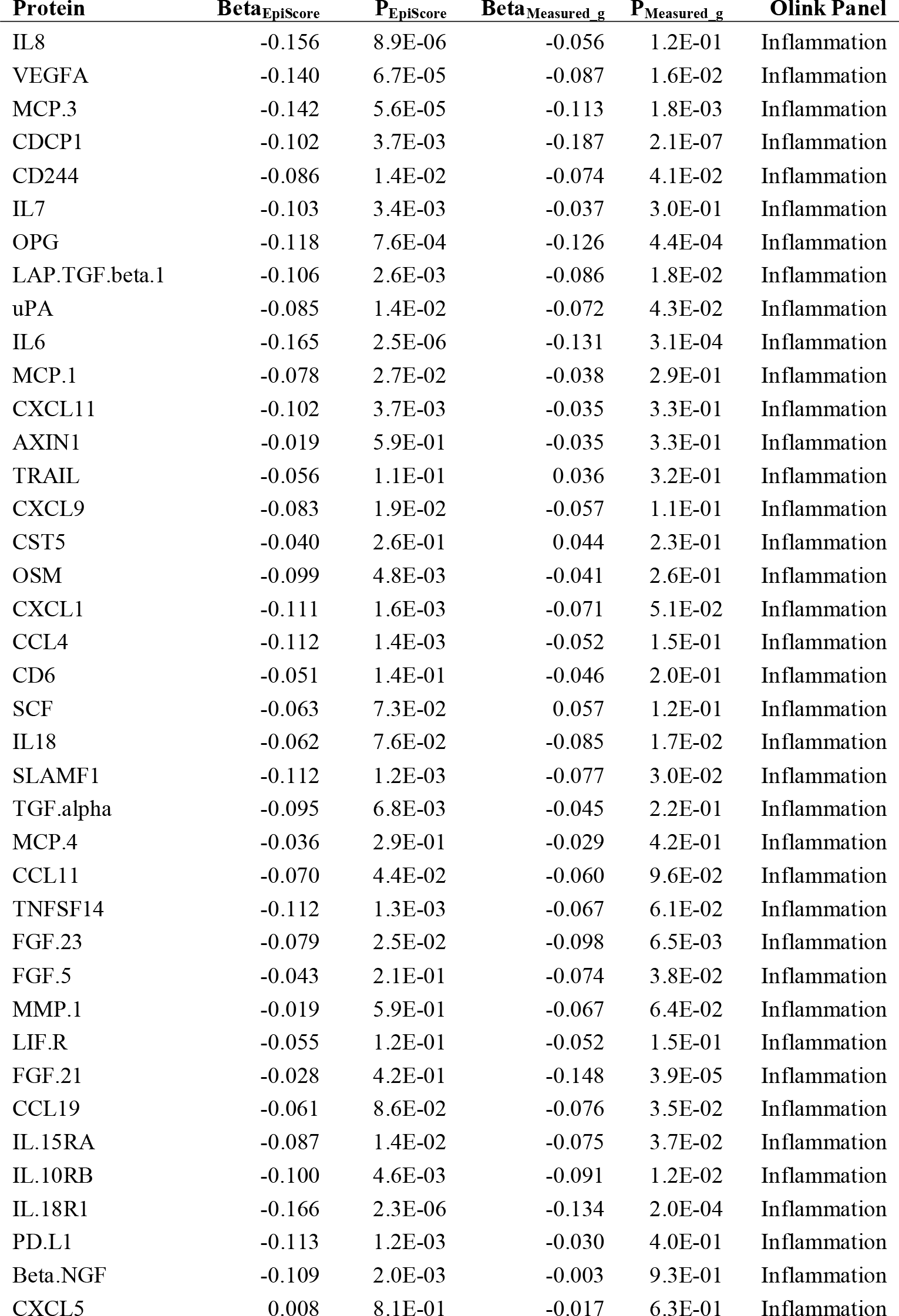
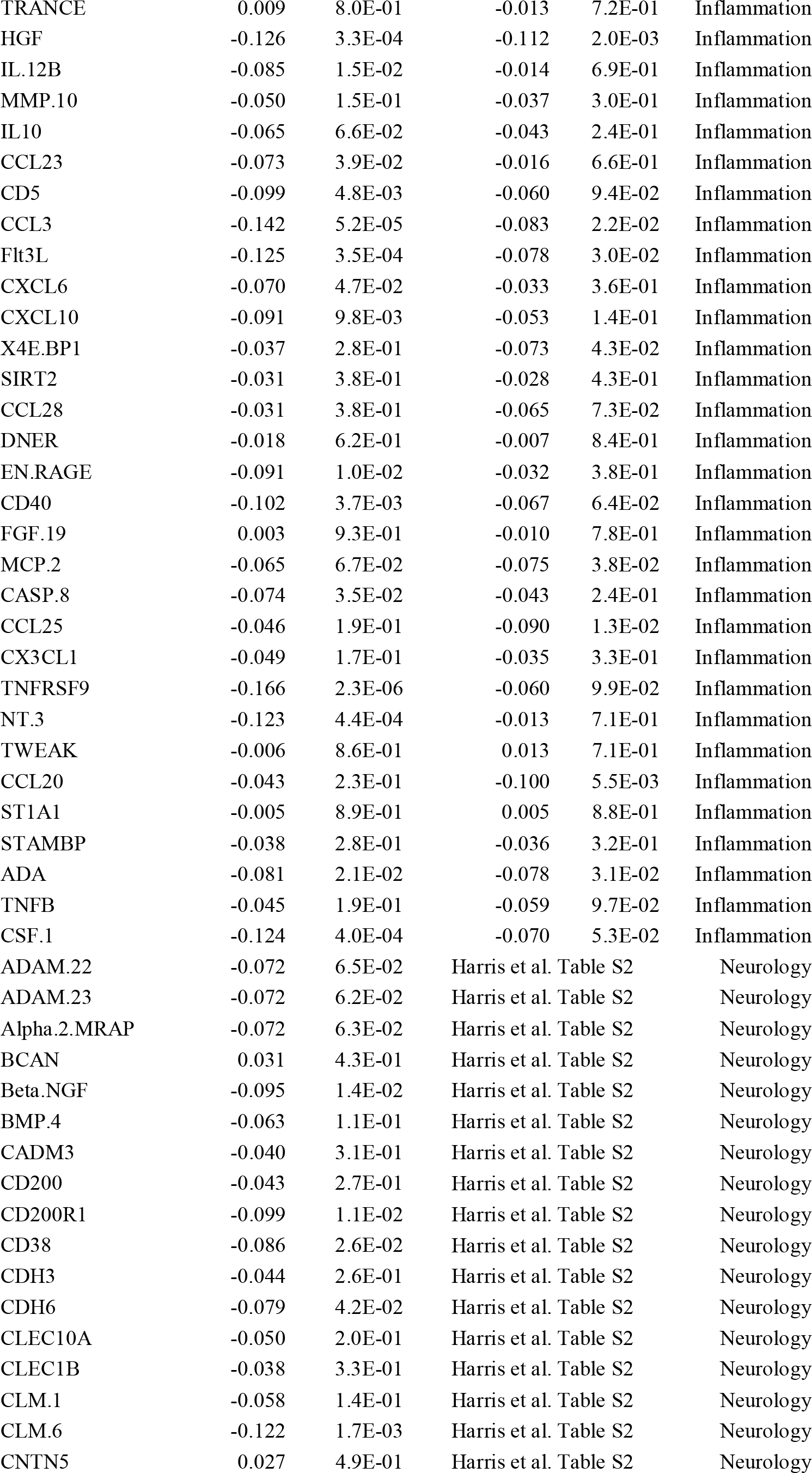
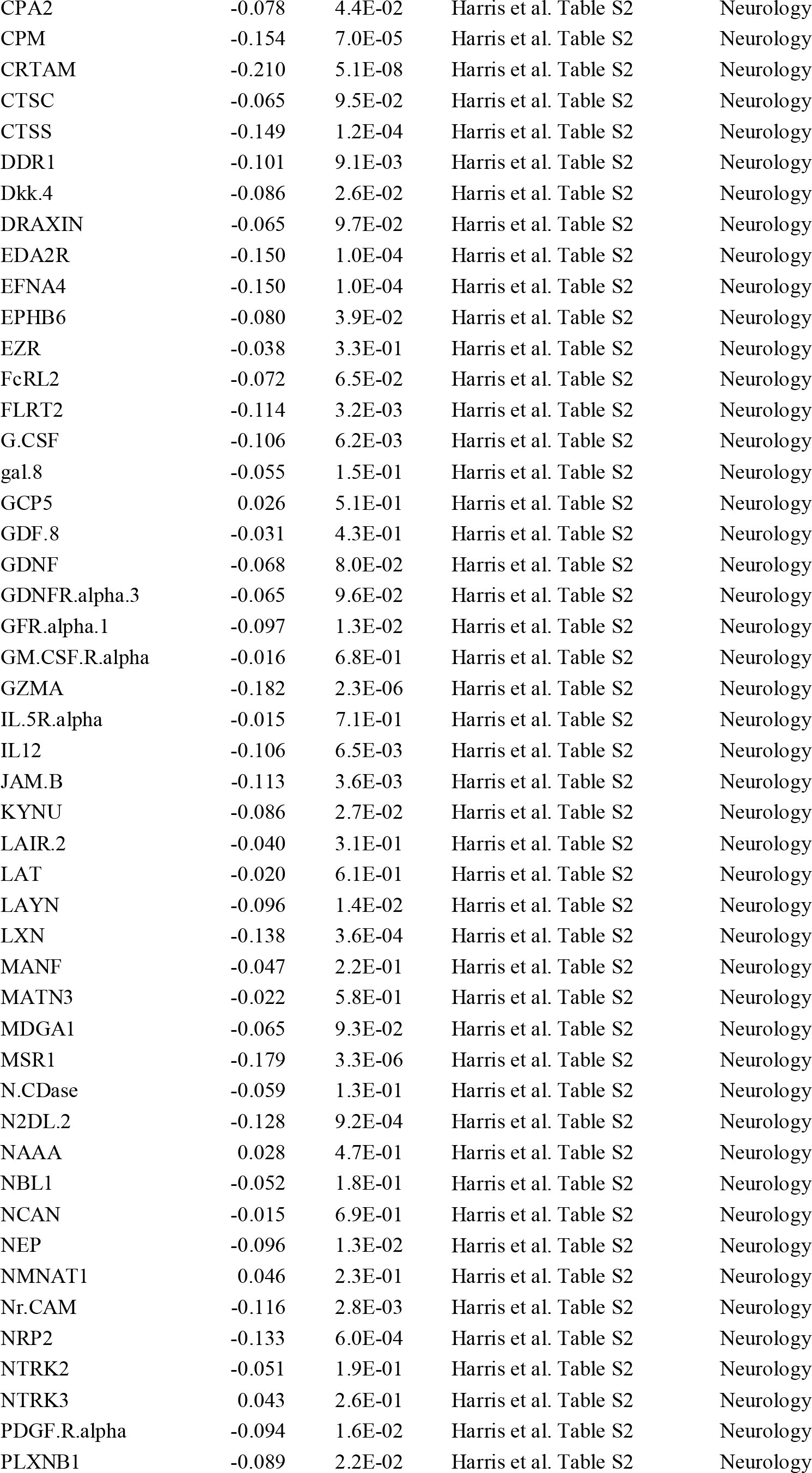
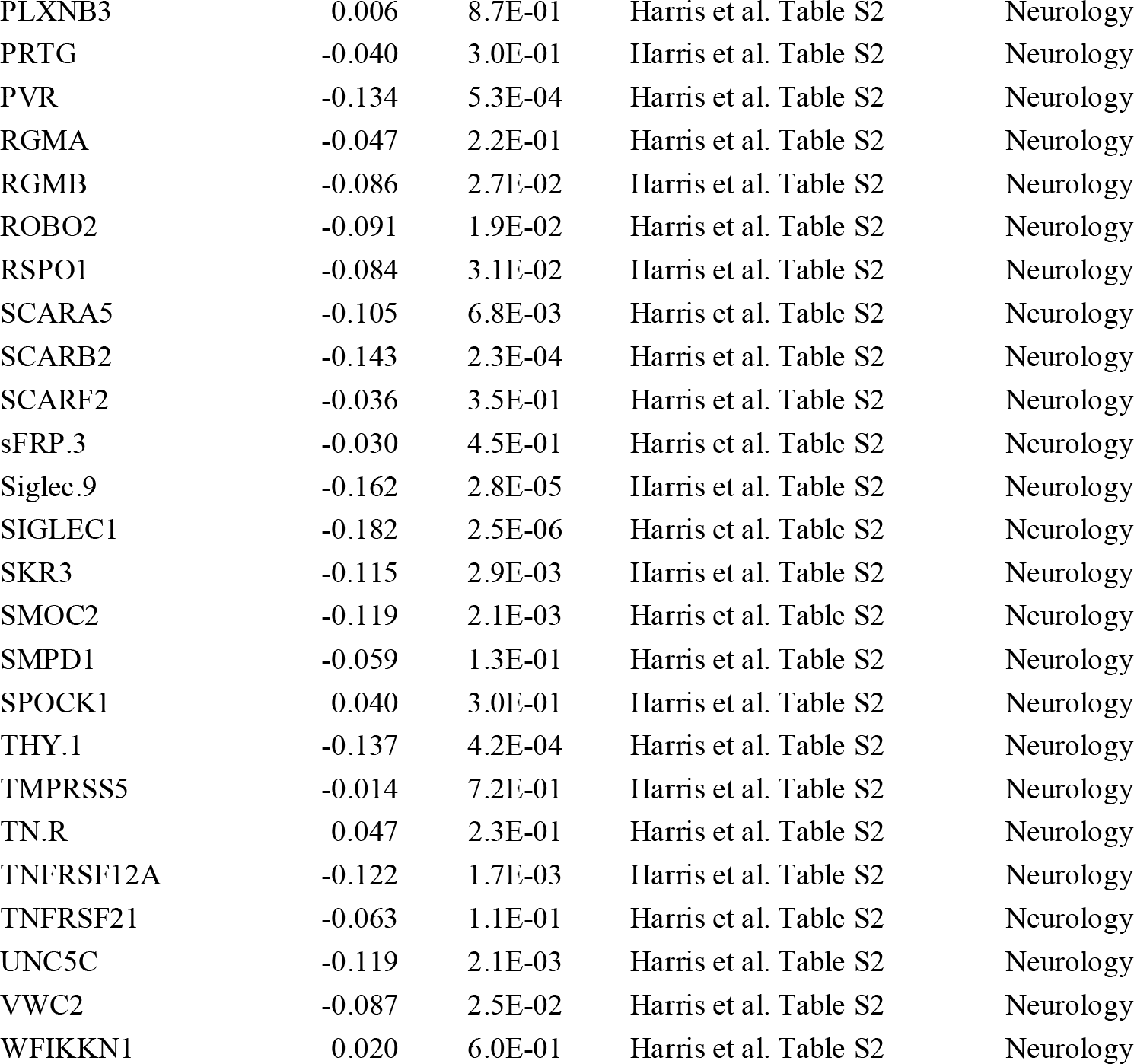
Age- and sex-adjusted linear regression associations between 70 inflammation-related and 90 neurology related proteins (outcomes) with the cognitive *g* Epigenetic Score (EpiScore) and measured cognitive ability in the Lothian Birth Cohort 1936. Neurology associations with measured fluid cognitive ability are reported in Supplementary Data 2 of Harris et al. Nature Communications.

**Supplementary Table 10.** Age at MRI scan- and sex-adjusted linear regression associations between the Epigenetic *g* Score (EpiScore) and measured *g* score and traits associated with global neuroimaging outcomes in the Lothian Birth Cohorts 1936. All continuous variables are standardised to mean 0 and variance 1.

| **Trait** | **$\beta_{\text{EpiScore}}$** | **$P_{\text{EpiScore}}$** | **$\beta_{\text{Measured } g}$** | **$P_{\text{Measured } g}$** |
| --- | --- | --- | --- | --- |
| Total brain volume | 0.15 | 3.8E-06 | 0.25 | 4.4E-14 |
| Grey matter volume | 0.14 | 4.3E-05 | 0.25 | 6.9E-14 |
| Normal appearing white matter volume | 0.15 | 1.3E-05 | 0.28 | 1.9E-15 |
| Global fractional anisotropy | 0.003 | 8.6E-01 | 0.11 | 4.6E-02 |
| Global mean diffusivity | 0.08 | 1.3E-01 | -0.08 | 1.2E-01 |

**Supplementary Figure 1:**
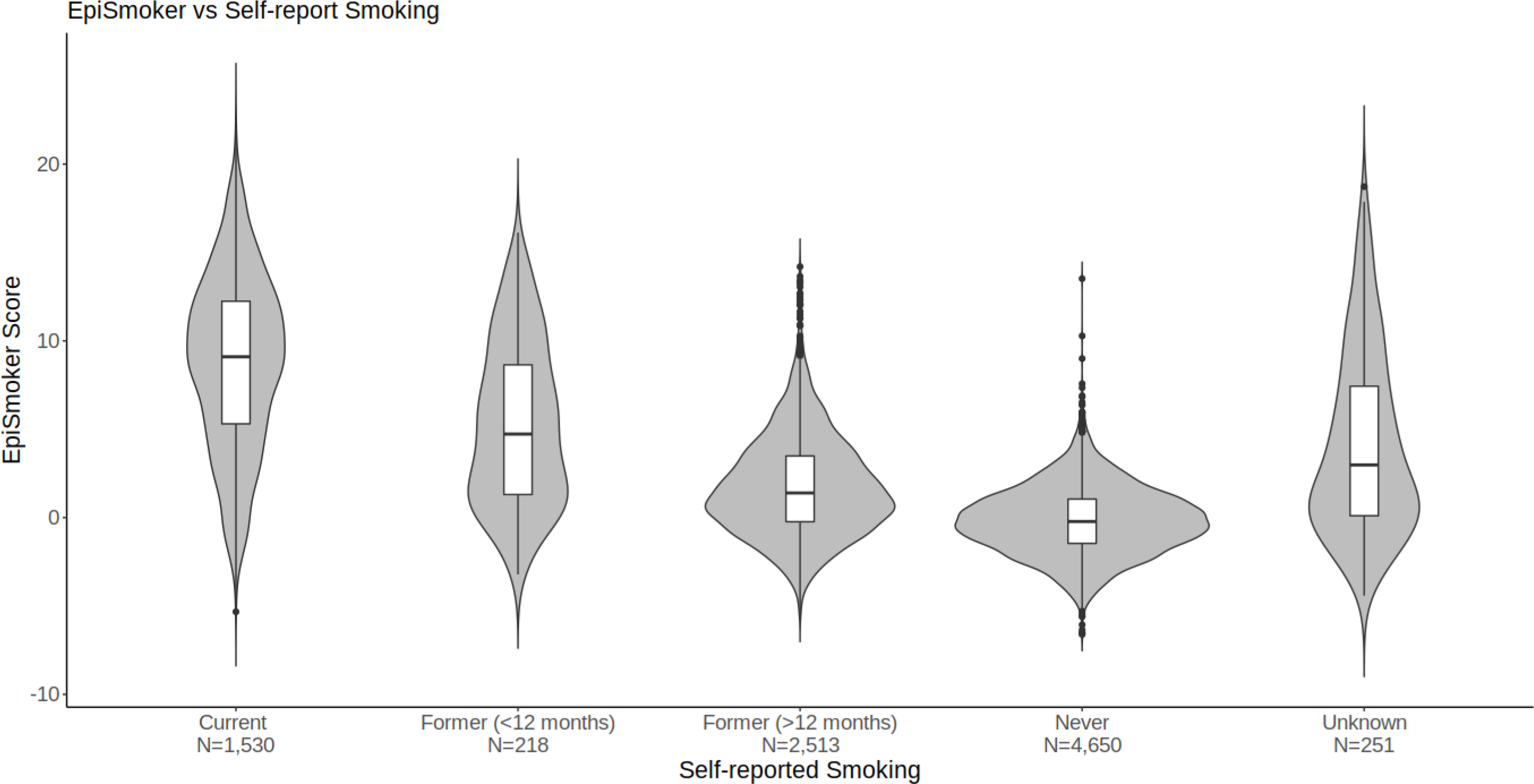
Plot of the epigenetic smoking variable (EpiSmoker) against self-reported smoking status (current, ever, never) in Generation Scotland (n=9,162).

**Supplementary Figure 2:**
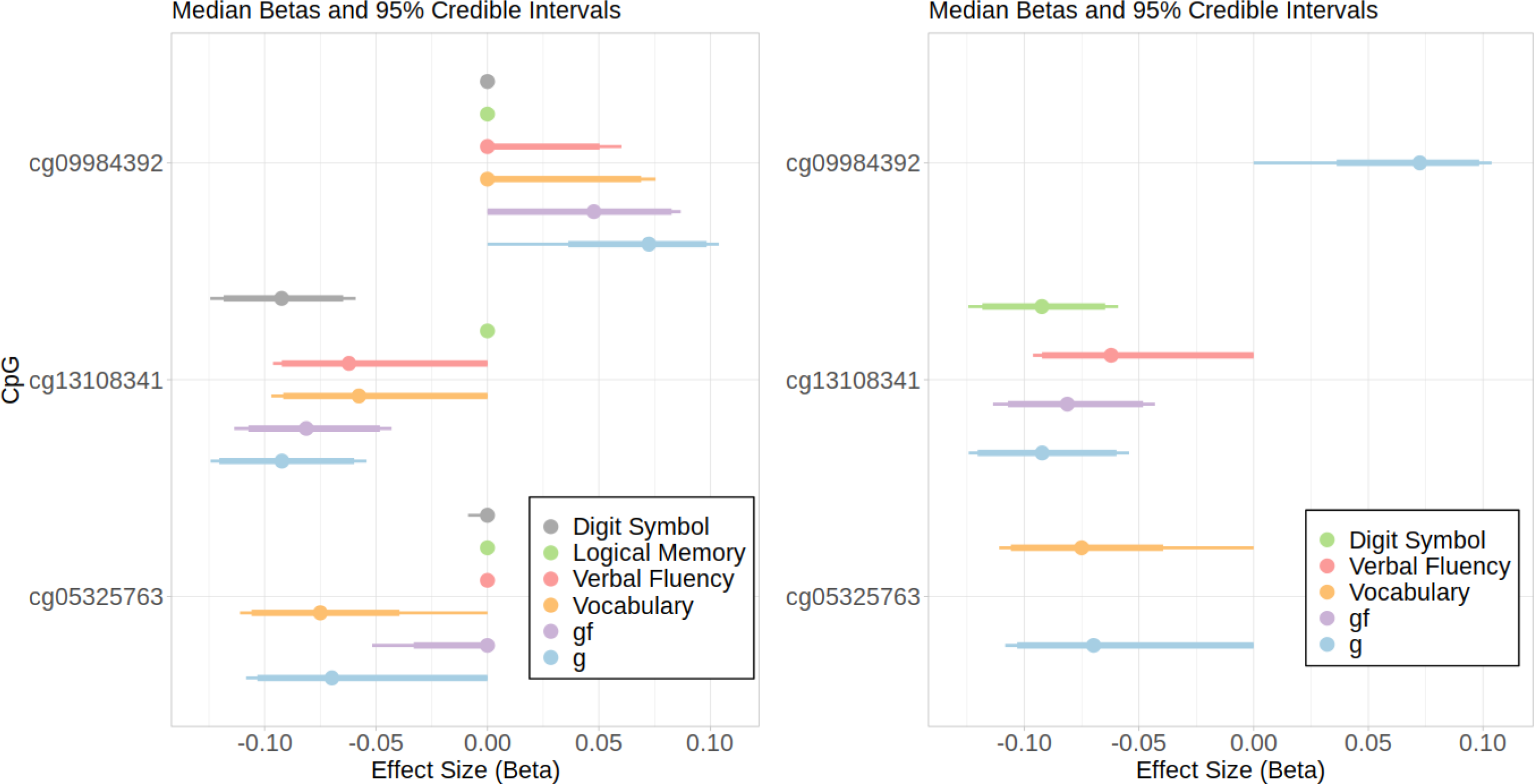
Median effects observed for DNA methylation probes with posterior inclusion probabilities (PIPs) > 0.8 across four cognitive tests and two composite measures (no probes identified for logical memory). Thick horizontal lines represent the 5^th^ and 95^th^ percentiles; thin lines represent 2.5^th^ and 97.5^th^ percentiles. Left panel displays summaries for all traits, right panel displays summaries for traits where PIP > 0.8.

**Supplementary Figure 3.**
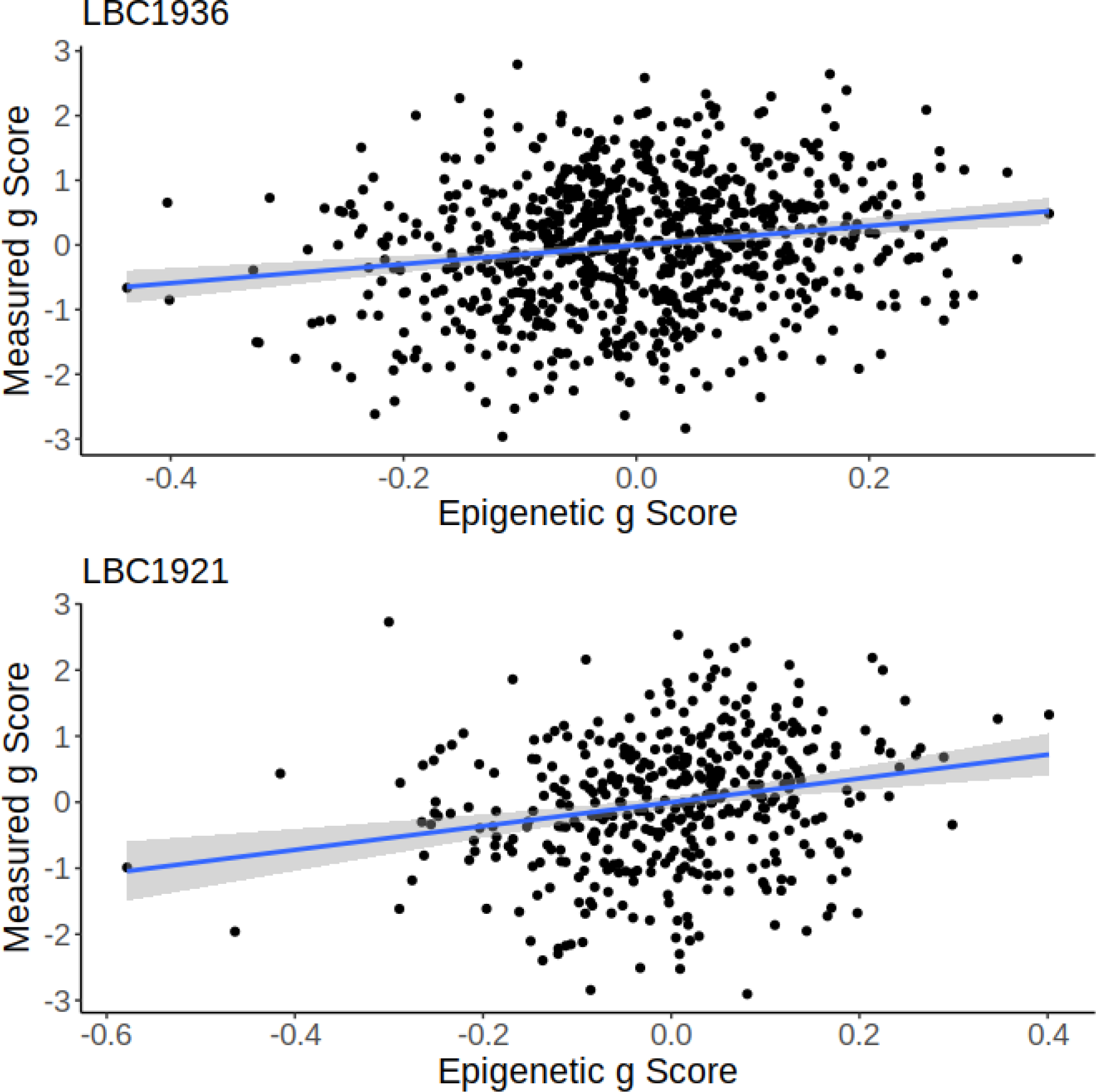
Scatter Plot of Epigenetic *g* Score by Measured *g* Score in the Lothian Birth Cohort 1936 (LBC1936) and the Lothian Birth Cohort 1921 (LBC1921).

**Supplementary Figure 4.**
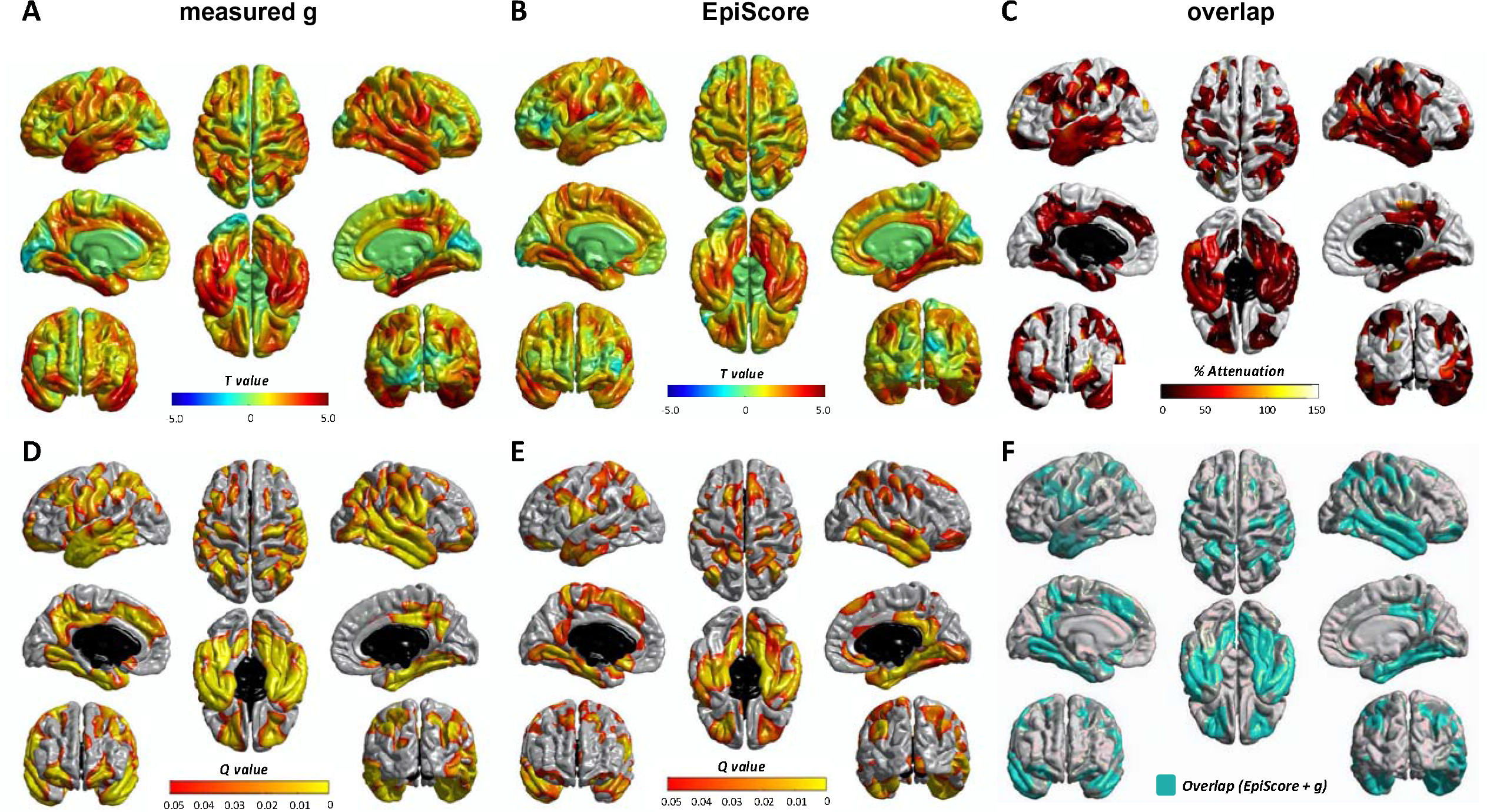
Regional cortical volume regressed against measured *g* (**left**) and EpiScore *g* (**middle**), colours denote the magnitude (T-maps; top, **A-B**) and significance (Q values; bottom, **D-E**) of the negative associations between cognitive measures and brain cortical volume. Panel (**C**) shows the percentage attenuation for the significant associations between EpiScore and cortical volume when also controlling for measured *g*. (**F**) shows the spatial extent overlap (green) in cortical loci that exhibit FDR-corrected unique associations.

**Supplementary Figure 5.**
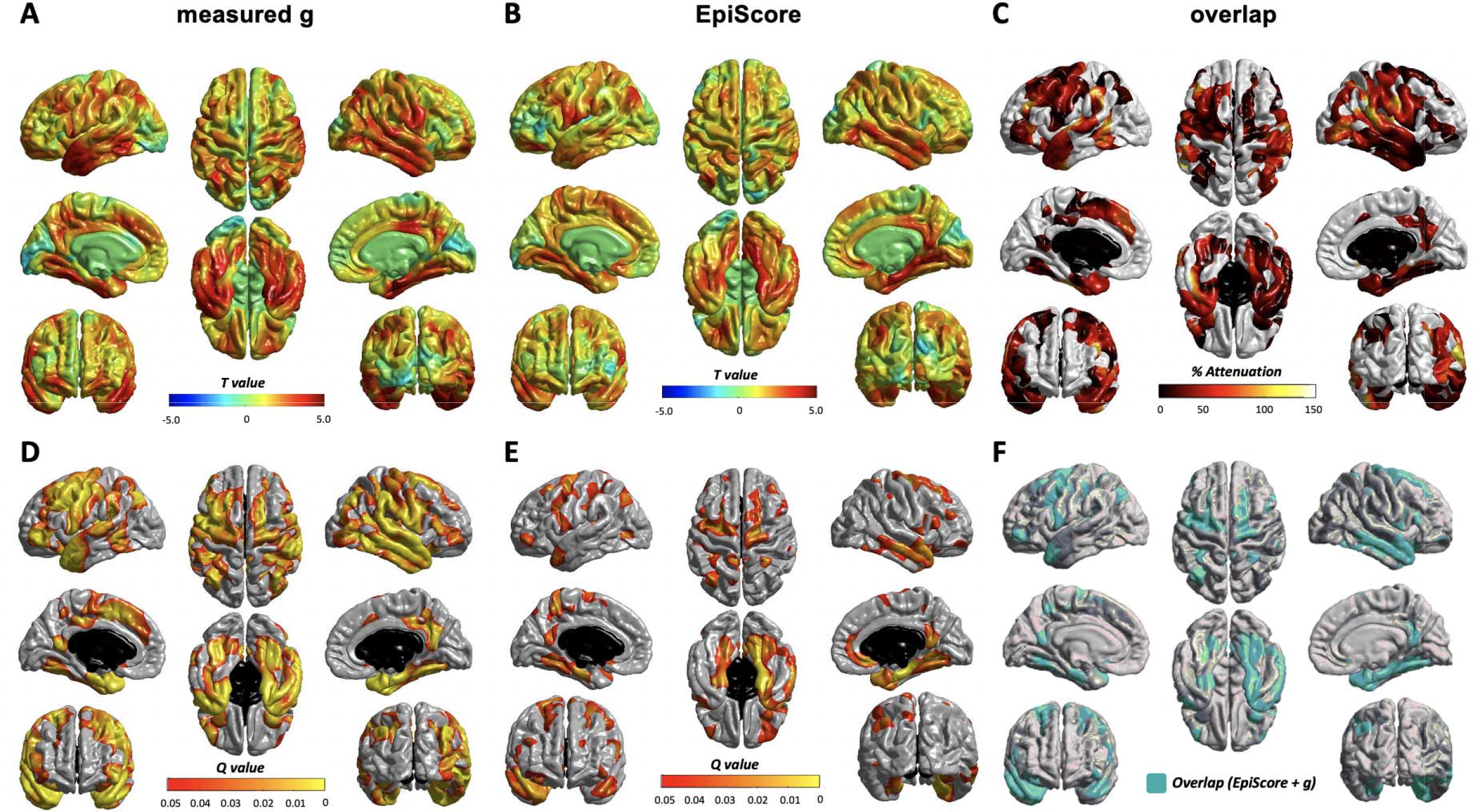
Regional cortical thickness regressed against measured g (**left**) and EpiScore *g* (**middle**), colours denote the magnitude (T-maps; top, **A-B**) and significance (Q values; bottom, **D-E**) of the negative associations between cognitive measures and brain cortical thickness. Panel (**C)** shows the percentage attenuation for the significant associations between EpiScore and cortical thickness when also controlling for measured *g*. (**F**) shows the spatial extent overlap (green) in cortical loci that exhibit FDR-corrected unique associations.

